# The Lactate Gradient, an unrecognized biomarker of potential significance

**DOI:** 10.1101/2022.09.21.22280074

**Authors:** Thomas Fabricius

## Abstract

**Background:** Human lactate formation is typically attributed to hypoxia, but it may actually be due to the influence of norepinephrine.

**Methods:** A lactate gradient was measured from the first few drops of blood obtained after application of a lancet to a finger pad. However, the blood needed to be carefully obtained according to this protocol.

**Results:** The measured lactate gradient was surprisingly large in some circumstances, and was not correlated with any of the other measured parameters such as oxygen or carbon dioxide blood levels. There were hints in the data that the lactate gradient seemed to be influenced by sympathetic noradrenergic nervous system activity.

**Conclusions:** The lactate gradient is relatively easily obtainable, and may reflect conditions causing enhanced sympathetic activity.

## Introduction

Lactate was first isolated from sour milk and was observed to be produced by lactobacilli. Lactate production was found to increase with cellular demand for ATP in the absence of sufficient oxygen. Due to these early studies, most medical practitioners have a conditioned belief that lactate formation is purely a waste product that begins and ends with insufficient oxygen. Hence, lactate has limited relevance in the current scope of practice, baring a few exceptions, such as a marker for sepsis in hospital care.

In research circles, lactate production and relevance has a much more nuanced appreciation. In the 1920’s, Warburg first reported on the high production of lactate *in the presence of adequate oxygen* in certain cancers. He called this process aerobic glycolysis (Warburg, 1925). As methods of biochemistry grew in sophistication, aerobic glycolysis was observed in a multitude of tissue types in mammalian cell lines. Positron emission tomography has shown that aerobic glycolysis even occurs in different brain regions in the setting of sufficient oxygen (Vaishnavi, 2010).

One way to promote aerobic glycolysis in tumors is through the effects of norepinephrine. This catecholamine is one of several secreted in response to sympathetic nervous system activity in the adrenal glands. Sympathetic stimulation was found to increase serum lactate concentrations in patients with sepsis (Jagan, et al. 2021). Wang et al (2021) found that norepinephrine enhances mRNA expression of several key glycolytic enzymes in tumor cells.

Besides the adrenal glands as a production source, norepinephrine is also a neurotransmitter and is released from sympathetic nerve terminals. Skeletal muscle has extensive sympathetic innervation. Norepinephrine will increase glucose uptake in skeletal muscle independent of insulin (Nonogaki, 2000). Increased sympathetic nerve activity promotes non-insulin mediated pathways of glucose uptake and utilization in muscle tissue (Nonogaki, 2000), likely through effects of norepinephrine.

The chemical kinetics of lactate formation are much faster than the kinetics of the Krebs cycle as the Krebs cycle has many more steps. The Krebs cycle is oxygen dependent and restricted to the mitochondria. Due to faster kinetics, lactate formation can quickly form ATP. However, aerobic glycolysis is less efficient than the Krebs cycle in overall ATP generation.

The sympathetic nervous system – norepinephrine – aerobic glycolysis pathway is amplified in oncology where chronic activation of the sympathetic nervous system is frequently observed (Nagaraja, 2013). Beta-adrenergic receptors on cell membranes are activated by catecholamines, including norepinephrine. Hence, beta blockers inhibit the effects of norepinephrine on these receptors, and interrupt the process of aerobic glycolysis in those tissues. An emerging cancer treatment adjunct is to add in a beta blocker to disrupt formation of a tumor’s preferred energy source, lactate, by inhibiting the activity of norepinephrine. This is an inexpensive and effective adjunct, with fewer side effects than traditional chemotherapy.

With this background, it should be apparent that the formation of lactate is far more nuanced than is typically appreciated in clinical medicine. It is also reasonable to understand that sympathetic innervation can promote glucose uptake and lactate formation in the tissue. It remains uncertain how best to measure tissue lactate from a practical standpoint, and what such readings might represent in both health and disease. One such method will be discussed.

## Materials and Methods

The participants were recruited from patients presenting to a Family Medicine clinic, and signed the informed consent form. The individuals were invited to participate in the study by submitting to two finger pokes to collect a few drops of blood, which were immediately analyzed. The first drop of blood was obtained with the subject sitting upright in a chair for anywhere from 5 to 25 minutes after arrival. The first drop of blood was obtained from using a Lactate Plus Meter Single-Use Safety Lancet, 28 gauge, 0.362mm x 1.8mm lancet applied to either the 2^nd^, 3^rd^, or 4^th^ fingers of the left hand. The first (and only) drop of blood was analyzed with the Lactate Plus Meter, and the value was recorded. The Lactate Plus meter required a sample size of 0.7 ml.

The subject was then asked to lie down on the exam table in a recumbent position on their back, with their right arm drooping downward off the edge of the table. This utilized gravity to bring more blood to the finger tip which assisted in collecting the 6 to 10 drops of blood in the capillary tube without resorting to squeezing the finger to force blood flow. The slightly larger BD Microtainer Contact-Activated Hi-Flow Lancet (1.5mm x 2.0 mm) was applied to the finger pad of one of the 2^nd^, 3^rd^, or 4^th^ fingers of the right hand. Between 6 and 10 drops of blood were collected in an EPOC Capillary Blood Collection tube, which holds 90 ml. Then, the last drop of blood from that finger was analyzed on the Lactate Plus Meter for comparison with the first drop collected as described earlier. The capillary tube was then immediately analyzed on the EPOC machine, and all of the values were recorded. Gauze was given and the subject applied light pressure to the lancet sites until homeostasis was quickly achieved. Figure 1.

**Figure 1.**
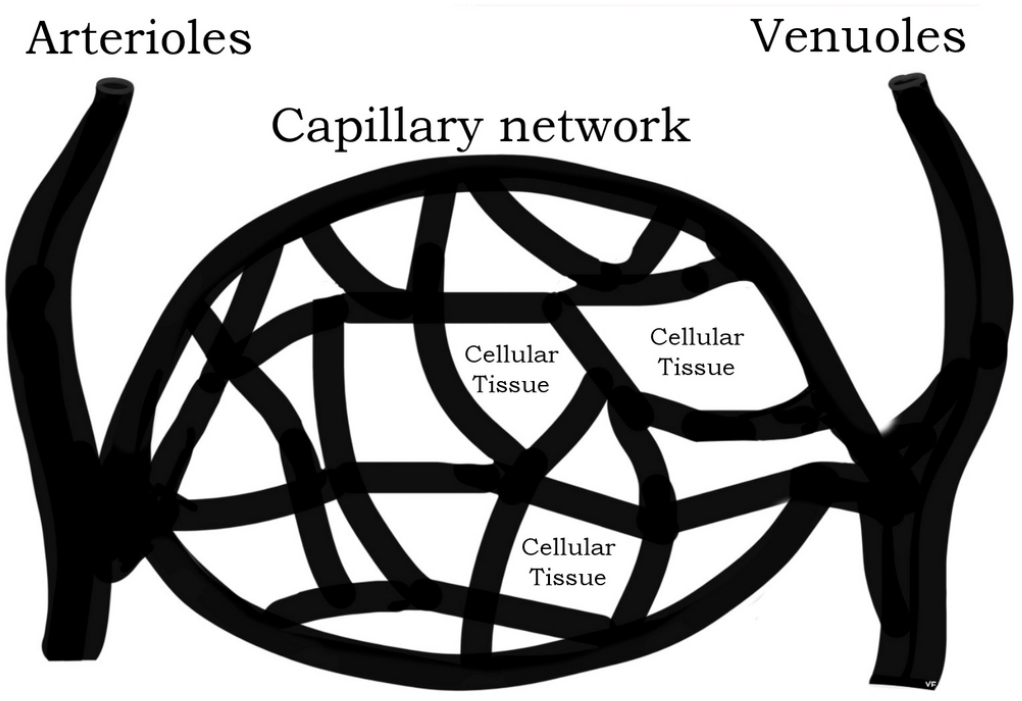
The smallest, shallowest lancet draws blood closest to the capillary networks, and therefore, the tissue itself. The volume of blood is very small. The larger, deeper lancet draws blood from the larger arterioles and venules, further away from the cellular tissues. Each successive drop of blood originates from deeper into the vascular system, reflecting chemistry closer to the central blood supply.

## Results

The most interesting result was the discrepancy between the two lactate values measured by the Lactate Plus meter. These two values were obtained from the first drop of blood from the shallow lancet applied to a left hand finger, and the last drop of blood collected from the slightly larger lancet blade applied to the right hand finger. Both drops were analyzed on the same Lactate Plus machine to exclude any effects of an unrecognized meter error. The 6 to 10 drops collected in the capillary tube analyzed with the EPOC analyzer would typically give lactate readings between the two readings from the Lactate Plus meter.

Figure 2.

**Figure 2.**
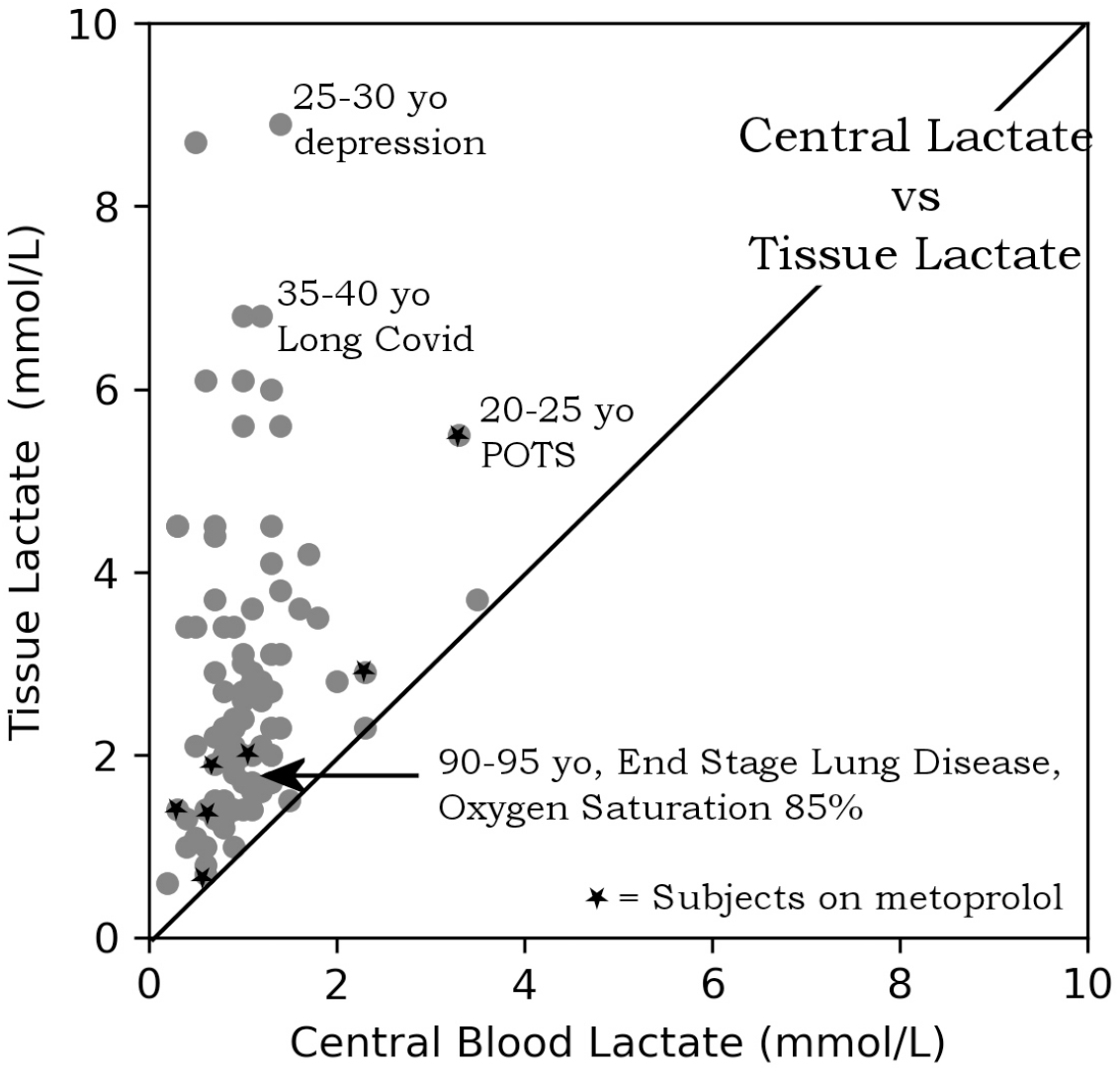
A plot of the lactate from the last drop of blood versus the first drop of blood collected for each test subject. Data points from participants taking metoprolol are labeled with a star. Beta blockers like metoprolol would be expected to block norepinephrine activity on cells resulting in lower lactate gradients.

**Figure 3.**
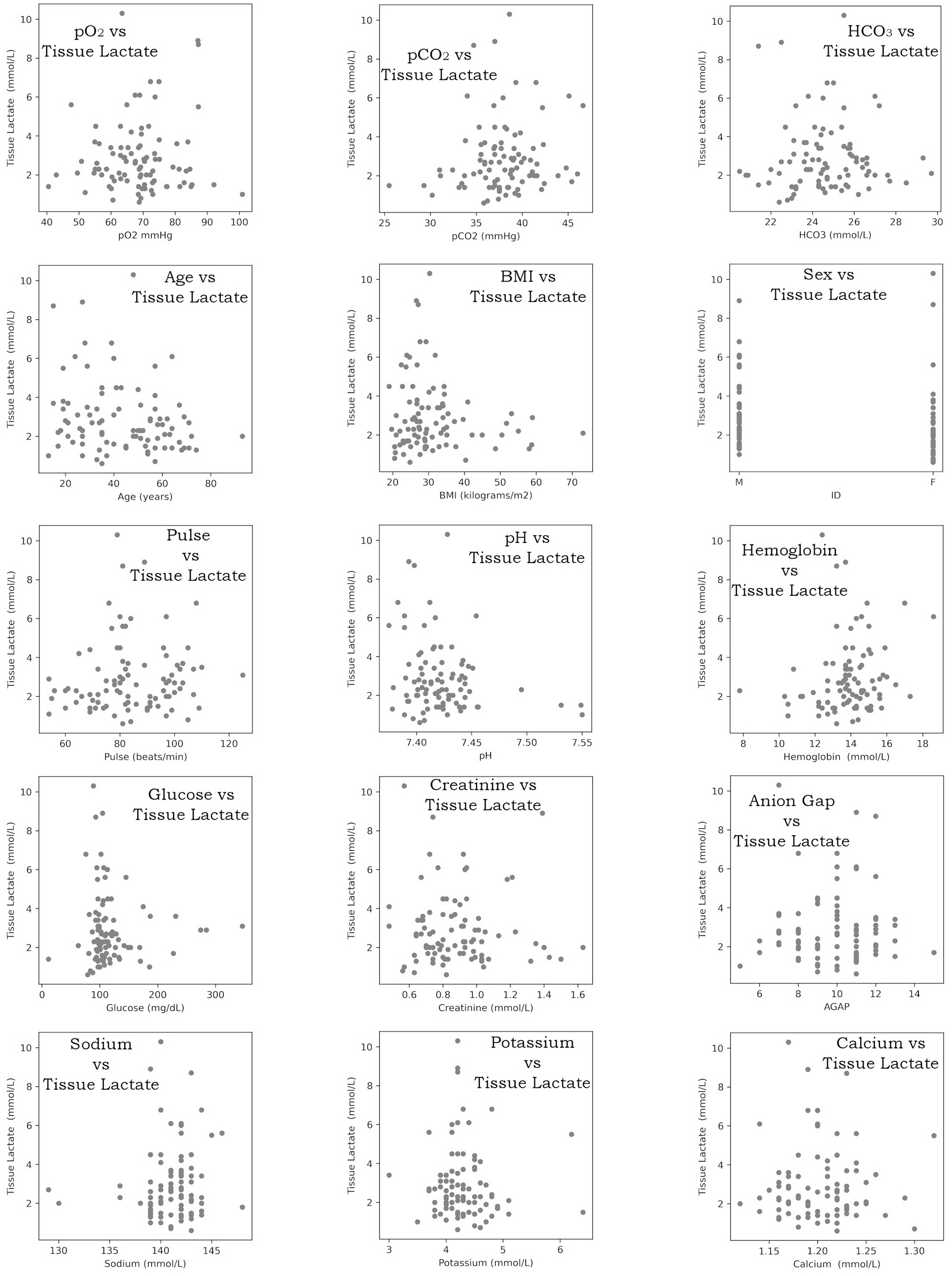
This is a collection of plots of tissue lactate versus various measured parameters. By visual inspection, there is no obvious correlation of tissue lactate with any of the measured parameters. Starting in the upper left, the measured parameters include: pO2, pCO2, HCO3, Age, BMI, sex, pulse, pH, hemoglobin, glucose, creatinine, anion gap, sodium, potassium, calcium.

Prior work has shown that the lactate readings from hand-held analyzers like the Lactate Plus meter correlate well with clinical grade lactate measurements (Bonaventura et al, 2015; Tanner et al, 2010). This was verified for the present study as three different Lactate Plus meters read lactate from the same blood samples consistent with each other. As the prior work found, the Lactate Plus readings correlated well with the EPOC reading, although they were slightly lower than the EPOC device readings. A linear correction factor could be applied if desired to bring all values into alignment. This can be found in the figure in the supplemental section. However, as the gradient was calculated between 2 readings from the Lactate Plus, the absolute value of the lactate reading wasn’t as important as the gradient.

The lactate reading from the first drop of blood will be referred to as the tissue lactate. The fact that the first drop of blood measured is still actually blood and not tissue only implies (but does not conclusively measure) higher tissue lactate levels. The last drop of blood collected and measured on the Lactate Plus meter will be referred to as central blood lactate, as the value more closely matches the blood lactate value in large arteries and veins.

This procedure identifies a lactate gradient in the blood between the smallest capillaries and the blood in the slightly larger arterioles and venules in the vascular tree. The presence of this gradient suggests that there can be significant differences in lactate throughout body tissues. The lactate gradient materialized in a volume of just 91.4 ml. The lactate gradient may be of significant diagnostic importance. Of the two readings comprising the lactate gradient, the tissue lactate measurement appears to carry the most importance as the central lactate typically falls in a much tighter range, and typically less than 2 mmol/L.

The tissue lactate levels did not correlate with any other measured parameters of the study. Specifically, no correlation was found between capillary pO_2_ and tissue lactate levels. Some of the highest tissue lactate readings were found in subjects with the highest pO_2_ levels. A wide variety of subjects were tested. One elderly (>90yo) participant with end stage lung disease and a very poor oxygen level of 85% had a relatively low tissue lactate level of 1.8. In comparison, some of the highest tissue lactate readings (>8) were found in physically healthy young adults, albeit with certain medical problems.

A review of the plots of age, sex, BMI, pulse, pO_2_, pCO_2_, hemoglobin, pH, bicarbonate, sodium, potassium, creatinine, calcium, hemoglobin, and anion gap did not show any distinct correlation with the tissue lactate reading. There was a rather large range in the size of the lactate gradient over the entire population studied.

## Discussion

The plot of tissue versus central blood lactate levels reflects the significance of the finding. Almost all of the central blood values were less than 2 mmol/L. This correlates well with the current hospital heuristic that lactate levels more than 2 mmol/L are a cause for concern and may indicate sepsis. As tissue lactate levels often exceeded 2 mmol/L, something significant is likely. The measurement of the lactate gradient is best observed when the volume of blood from the first reading (from the shallowest lancet) is as small as possible (0.7 ml). It would appear that the blood has compensatory mechanisms to clear lactate, likely to prevent acidosis. The comparatively larger volume of blood required for the EPOC device to measure lactate would completely obliterate measurement of the gradient by itself.

There is no discussion in the literature regarding prior observation of the lactate gradient. However, the literature review in the introduction provides some obvious hints at what may be causing this gradient formation.

The central versus tissue blood lactate plot is labeled with some subject diagnoses. It is readily apparent that some subjects with mental health diagnoses have much higher lactate gradients, regardless of age. However, not everyone with a mental health diagnosis had an elevated tissue lactate. Anecdotal factors like medication effects or time since the last meal might play a role. This may be expected as fasting allows the body to clear lactate from the central blood supply through the Cori cycle. Fasting also promotes an increase in parasympathetic tone. Thus, someone with depression who hasn’t eaten in the last 18 hours may have a lower lactate gradient. The author has seen a strong association between fasting and lower tissue lactate in periodic self-testing.

There are signals in the data that certain medications play an important role in the measured lactate gradient. As would be expected, beta blockers, and some mental health medications like anti-psychotics, seem to have some correlation in lowering the lactate gradient. The circulation of blood will continuously wash out the the capillary (tissue) lactate level. The rate of lactate replenishment would be expected to reflect the current activity of the sympathetic nervous system in that tissue at that time.

The location of blood collection also gives some hint as to the possible genesis of the lactate gradient. High performance endurance athletes test their blood to determine their lactate threshold for training. Athletes utilize a drop of blood from their ear lobe as a more reliable indicator of their central lactate level (Raa et al, 2020). The finger pad was chosen as the site of blood for collection for this study as the finger(s)/hand have a much higher concentration of sympathetic innervation in the dermis than is found at other areas of skin. Thus, there are multiple lines of preliminary evidence that the lactate gradient partly reflects the activity of the sympathetic nervous system.

Typically, measurement of autonomic nervous system activity is time consuming, expensive, burdensome, and is confined to specialty autonomic laboratories. The idea that the current state of the sympathetic noradrenergic nervous system activity may be partially available through a very inexpensive point-of-care finger-poke blood test has tremendous appeal. A companion paper will soon be released showing how the lactate gradient can be provoked with maneuvers that activate the sympathetic nervous system. The caveat is that the lactate gradient reflects the sum total of many different influences on autonomic activity, including circadian rhythms, timing and composition of last meal, medication, concurrent illness, illicit substances, menstrual cycle, life stresses from work and relationships, activity level, lunar cycle, weather pattern, and many others not presently listed. It is not feasibly possible to control for all influences on autonomic activity.

## Conclusions

The author present a new measurement of a previously unrecognized lactate gradient that can be obtained at the bedside. There is some cautious optimism that the gradient can shed light on the current activity of the sympathetic noradrenergic nervous system. The idea that cellular level metabolism affecting lactate production can be quickly influenced by the nervous system in a tissue-specific manner is an enticing thought. This can point to a potentially new understanding of many different disease processes through a lens of lactate. The primary care population in this study often had presenting complaints including irritable bowel, chest pain, palpitations, anxiety, fibromyalgia, fatigue, and even the elusive Long Covid. If the sympathetic noradrenergic influence on the lactate gradient can be duplicated and confirmed, then measurement of the lactate gradient may help clinicians recognize when patient somatic and psychiatric complaints may be due in part to autonomic states of high sympathetic activity. These findings must be further explored, replicated, and refined to achieve that goal. This study only describes a novel measurement of a potentially clinically relevant marker. It has not provided definitive endpoints of clinical interpretation of the lactate gradient.

## Data Availability

All data produced in the present work are contained in the manuscript.

## Institutional Review Board Statement

This study was approved by the SSM Agnesian Institutional Review Board.

---

**Figure Supplemental Section.**
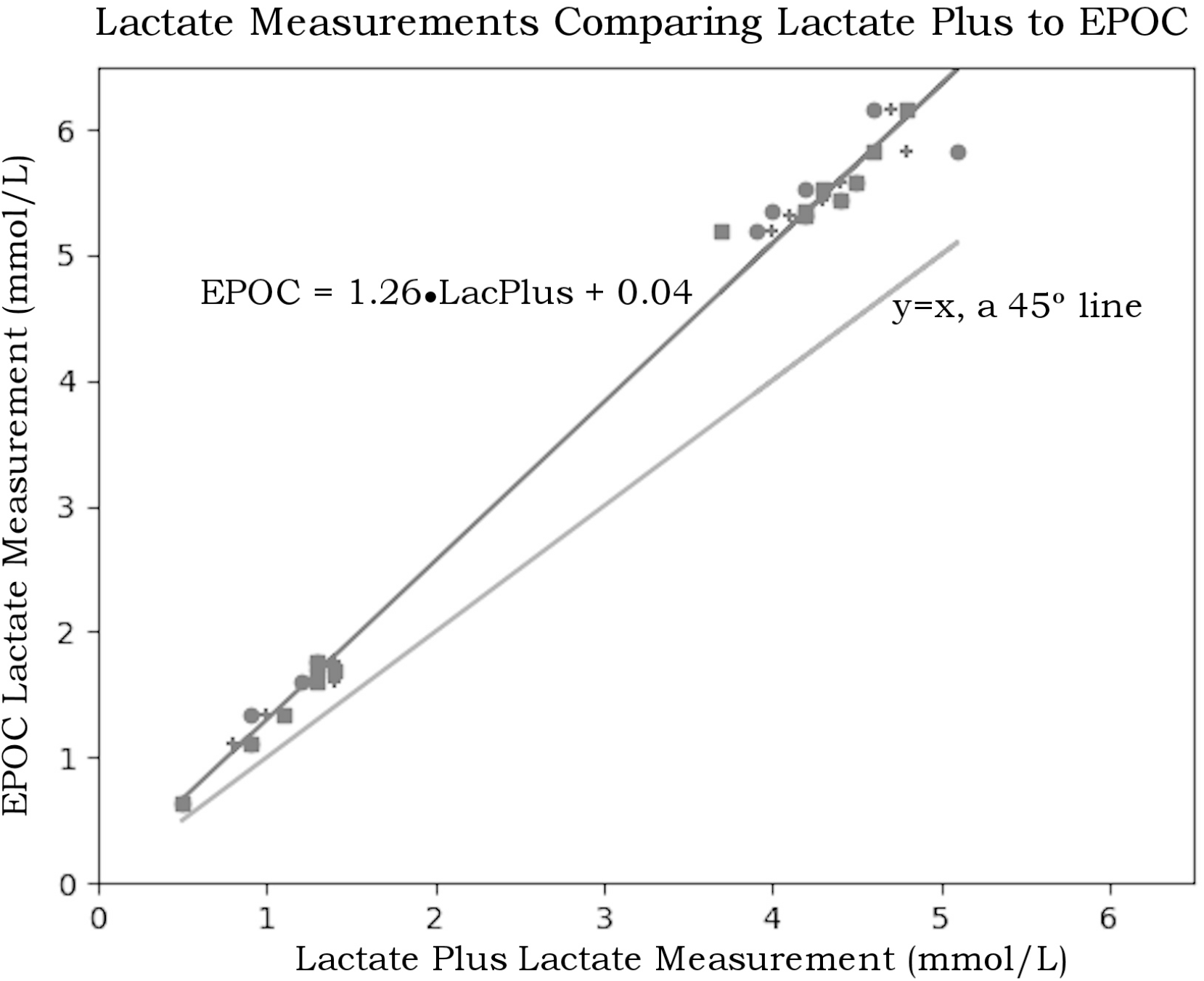
From a central blood sample, three Lactate Plus meters measured consistent to each other, but measured lower lactate readings compared to the EPOC device.

